## Supplementary material for "Opportunities to sustain a multi-country quality of care network: lessons on the actions of four countries Bangladesh, Ethiopia, Malawi, and Uganda": S1: Text. PLOS Global Public Health QCN Evaluation Collection 2-page summary.

**PLOS Global Public Health QCN Evaluation Collection – Introducing the 9 papers and how they fit together**

Our collection of 9 papers are the main results from our **QCN Evaluation** project evaluating the [WHO-coordinated Network for Improving Quality of Care for Maternal, Newborn and Child Health](http://www.qualityofcarenetwork.org/stakeholder-and-community-engagement-quality-care-initiatives) (Quality of Care Network, or QCN). QCN is supported by three UN agencies (WHO, UNICEF & UNFPA) and major global funders USAID, BMGF, and others. Together they mobilised millions of dollars in direct financing or in-kind technical support, and QCN is a 'Flagship' programme. Given this level of investment and the growing interest in networks to facilitate health system development it’s evaluation is crucial. The full title of our research project describes the overall question we set out to answer:  *How does a multi-country, multilateral network focused on specific health care improvements evolve and what shapes its ability to achieve its goals?* We hope our collection succeeds in providing some useful answers. Our collection includes results from our work evaluating the network at the global level and at national and local levels in each of our four case study countries: Bangladesh, Ethiopia, Malawi and Uganda. Country-specific papers will also be forthcoming outside of this collection.

Our research was conducted over three years during 2019-2022 and involved a series of iterative rounds of mixed methods data collection including stakeholder interviews at global, national and local levels; observations of best and least performing facilities as well as QCN international and national meetings; review of key documents; and surveys of QCN members covering their perception of the work of the network, and interactions between them.

We asked three broad questions concerning the emergence, legitimacy and effectiveness of the network (Box 1) and these are tackled directly in turn in our papers by Shawar *et al* [Paper 1], Akhtar *et al* [Paper 2] and Djellouli *et al* [Paper 3]. Shawar *et al’s* paper [Paper 1] focuses on factors shaping emergence of the network at global and national levels and finds a spectrum of emergence across our four case study countries. Akhtar *et al* [Paper 2] look at interactions between institutions within each of our four countries, and with the global level to shed light on the ownership of the work of the network and its legitimacy from the perspective of national governments. The extent of network emergence, ownership and legitimacy has a bearing on the effectiveness of the network, which is then investigated in depth in the paper by Djellouli *et al* [Paper 3]. This paper investigates, from global to national and local levels, the policy process, operationalisation of the network, delivery of locally valued interventions and effectiveness of the network in changing processes that should improve maternal, newborn and child health outcomes.

**Box 1: QCN Evaluation research questions:**

**1. Network emergence**: What attributes of this multi-country network and its operational strategy and performance affect the engagement of network actors at global and national levels and their adoption of a shared agenda and goals to improve maternal and newborn health services?

**2. Network legitimacy and ownership**: What shapes the relationship between country teams and the global network leadership and how does this influence ownership of the policy and management work that is required to set national aims and improve services, and which characteristics of the health system context appear to influence this?

**3. Network effectiveness:** What specific form does national QCN activity take and how does this influence which specific interventions are delivered, which of these are felt to be successful by local actors and which lead to measurable changes in processes and outcomes.

The other papers then cover key areas of our work in detail, also contributing to answering our research questions. The paper by Mukinda *et al* [Paper 4] is on our quantitative stakeholder network analysis to explore and describe the structure of the QCN and map actors at different levels by examining the frequency and quality of interactions between them. It is relevant to both research question 1 on network emergence and research question 3 on network effectiveness: via quantifying the frequency and quality of interactions between individuals and different types of stakeholders, it shows the extent of network emergence and how network effectiveness may be mediated by such interactions. The paper by Nkhata *et al* [Paper 5] develops a theory of change of how the network has been operating in practice to achieve the results presented in the other papers, and contrasts this to the initial theory of change proposed by the WHO and partners at the inception of the QCN. It therefore aids our understanding of processes that contributed to network effectiveness (or lack of it).

Tesfa *et al* [Paper 6] investigate individual, organisational and system capacities enabling or hindering the functioning of the network. These results are useful for understanding both QCN emergence and effectiveness, and also have a bearing on network legitimacy. Mwandira *et al’s* paper [Paper 7] is concerned with network effectiveness and looks specifically at how well the QCN has been able to facilitate innovation, learning and sharing within and between countries, presenting examples at local, national and global levels. The paper by Lemma *et al* [Paper 9] then looks ahead considering actions taken in our four case study countries that may create opportunities to enable, or hinder the sustainability of the work of the QCN. Finally, Seruwagi et al [Paper 8] reflect on our experiences of evaluating the QCN over the past three years, drawing lessons that we hope may be useful for others undertaking similar research in future.

To streamline our papers, avoid overlap of material, and improve integration and coherence of the collection, our papers share two common supplementary material files: a common methods document [ref: ‘QCN papers common methods section.docx’] covering all of the methods and methodology used in our QCN evaluation project, and a common background and context document [ref: ‘QCN papers country context supplement.docx’] containing key background information on our four case study countries. We suggest these are read after reading this introduction. Individual papers in our collection may reproduce or summarise different specific components of these documents as required for their specific purposes.

Several cross-cutting issues are covered in our collection, but from different and complementary angles across the different papers. Barriers and facilitators to the operation of the QCN are covered in detail in four papers, though Shawar *et al* [Paper 1] focus on the beginning of the network (its emergence), Djellouli *et al* [Paper 3] and Tesfa *et al* [Paper 6] focus on the middle – its operation and effectiveness during the time of study: 2019-2022, and Lemma *et al* [Paper 9] focus on the future (sustainability) of the network. Both the papers by Akhtar *et al* [Paper 2] and Mukinda *et al* [Paper 4] look at interactions between QCN stakeholders in depth, though Akhtar *et al* [Paper 2] develop a qualitative understanding of different kinds of institutional interactions, whilst Mukinda *et al* [Paper 4] present a quantitative mapping of individual level interactions, grouped into types. The effectiveness of the QCN is discussed across three papers in this collection. Djellouli *et al* [Paper 3] adopt a wide lens looking at network outputs, policy consequences and impacts at all levels of governance, while Mwandira *et al* [Paper 7] investigate in more depth the effectiveness of the network in terms of innovation, learning and sharing and Nkhata *et al* [Paper 5] concentrate on comparing the initial theory of change with the actual operationalisation of the network.

We provide a summary synthesis of the key findings across our nine papers in a separate accompanying commentary.[ref] We hope you enjoy reading our collection and that it offers findings that are useful to future multi-country networks focused on implementation.
