## Supplementary material for "Opportunities to sustain a multi-country quality of care network: lessons on the actions of four countries Bangladesh, Ethiopia, Malawi, and Uganda": S3: Table. Data coding and analysis matrix

| **Themes and codes under each sustainability action points** | | | | | |
| --- | --- | --- | --- | --- | --- |
| **Planning opportunities for reflection and adaptation** | **Government ownership and Transition of responsibilities** | **Motivating micro-level actors** | **Institutionalization within the health system** | **managing financial uncertainties** | **Fostering community ownership and acceptance** |
| **Network Effectiveness:** network emergence, | **Intervention function:** enablement, Challenges to network effectiveness | **Intervention function:** Incentivisation, | **Intervention function:** environmental restructuring, enablement, incentivization, Training, Capacity | **Policy categories:** Fiscal, guideline | **QCN implementation in country** |
| Network organisation and effectiveness | **Policy categories:** Fiscal, guideline, legislation, Political stability | **Factors influencing adoption of innovation** | **Factors influencing adoption of innovation:** compatibility, complexity, observability, relative advantage, tribality | **Conditions in the global policy environment:** Funding | **LALA:** Community Engagement/Accountability, |
| **Network implementation:** LALA, | **Features of the network actors:** governance arrangement and coordination, leadership | Enablement | Diffusion of innovation - communication channels |  |  |
|  | **Network Cohesion:** Coherence, collective participation | QCN implementation in country | Network accomplishments |  |  |
|  | **Network implementation:** Network accomplishments | Resources for network activities | LALA |  |  |
|  | **Network implementation:** LALA | Motivation | QCN implementation in country |  |  |
|  | **Network implementation:** QCN implementation in country | Intervention function: Incentivisation, | Policy categories: Service Provision, Communication marketing |  |  |
|  | **Network implementation:** Resources for network activities |  | Case Piloting and scaling up |  |  |
|  | **Case** **Contextual Factors:** Existing MNCH policies, Health System |  | Case Outcomes in Bangladesh |  |  |
